# C-reactive protein and high-sensitivity C-reactive protein levels in asymptomatic intestinal parasite carriers from urban and rural areas of Gabon

**DOI:** 10.1101/2023.04.08.23288297

**Authors:** Helena Noéline Kono, Mérédith Flore Ada Mengome, Bedrich Pongui Ngondza, Roger Hadry Sibi Matotou, Luccheri Ndong Akomezoghe, Bernadette Ekomi, Bridy Chesly Moutombi Ditombi, Jeanne Vanessa Koumba Lengongo, Jacques Mari Ndong Ngomo, Noé Patrick M’Bondoukwé, Cyrille Bisseye, Denise Patricia Mawili-Mboumba, Marielle Karine Bouyou Akotet

## Abstract

**Background:** Chronic carriage of intestinal parasitic infections (IPIs) can induce chronic inflammation and dysbiosis, which are risk factors for non-communicable diseases. The objective of this study was to determine the relationship between IPI carriage and inflammation in a population of volunteers living in Gabon.

**Methodology and Principal Findings:** A cross-sectional study was conducted from September 2020 to November 2021 in asymptomatic participants aged 18 years and over residing in different areas of Gabon: Libreville (urban area) and Koula-Moutou and Bitam (rural areas). The detection of IPIs was carried out using common techniques. Inflammation markers, C-reactive protein (CRP), and high-sensitivity C-reactive protein (hsCRP) were assayed. Overall, 518 participants were included, 64.5% (n = 334) of whom resided in urban area and 35.5% (n = 184) in rural areas. The median age was 35 years [27; 46]. The prevalence of asymptomatic IPIs was 29.9% (n = 155), with a significantly higher frequency in rural areas than in urban area (adjusted OR 6.6 [CI 3.2-13.8], *p* < 0.01). Protozoa were more frequent than soil-transmitted helminths (STHs) in both areas: 81.6% (n = 40) in urban area and 69.8% (n = 74) in rural areas. STHs were predominant in rural areas (48.1%) than in urban area (22.4%). High concentrations of hsCRP and CRP were significantly more frequent in inhabitants of rural areas (23.4% (n = 43) and 56.5% (n = 104), respectively (*p* < 0.01) than those of urban area (11.1% (n = 37) and 34.5% (n = 116), respectively; *p*<0.01). High levels were more frequent in parasitized individuals (for hsCRP, 22.6%, n = 35, *p* < 0.01, for CRP, 52.9%, n = 82; *p* < 0.01); in particular among STH carriers (for hsCRP; 65.9%, n = 27, *p <* 0.01, for CRP: 36.6%, n = 15; *p* < 0.01).

**Conclusions/Significance:** This first study showed that asymptomatic IPIs, especially STHs, are associated with higher CRP and hsCRP levels. Others biomarkers of inflammation must be analyzed to confirm the relationship between asymptomatic IPIs and chronic inflammation.

**Author summary:** Repeated or chronic parasitism can maintain local or systemic chronic inflammation, CRP and hsCRP are sensitive biomarkers of subclinical low-grade inflammation. This study assessed the serum levels of CRP and hsCRP in adults with or without intestinal parasite (IPI) carriage according to residence area and parasite species. IPI chronic carriage, especially with pathogenic protozoa and/or STH, was associated with higher levels of CRP and hsCRP. These findings suggest that intestinal parasite carriage contributes to low grade systemic inflammation which is a driver of NCD. The role of chronic carriage of other enteropathogens on chronic inflammation status as well the relationship between IPI and dysbiosis should be further analyzed in endemic countries.

## Introduction

Intestinal parasitic infections (IPIs) due to intestinal protozoa and helminths including soil-transmitted helminths (STHs) still constitute a public health problem in sub-Saharan Africa (SSA). The IPI burden is due to favorable climatic conditions, the absence or insufficient hygiene and sanitary measures, and poverty [1]. In endemic areas, IPIs are often more frequent in rural than in urban areas, and are generally asymptomatic and thus undiagnosed, leading to chronic parasite carriage [1].

Moreover, while diet and smoking are more frequently associated with the occurrence of non-communicable diseases (NCD) in high-income countries, with NCD-related mortality associated with older age groups, other factors such as younger age, could be a cause in low- and middle-income countries (LMIC) where NCD related deaths frequently occur in 30-69-year-old individuals [2]. Therefore, it is important to determine region-specific modifiable NCD risk factors. Apart from diet and smoking for example, environmental factors and infectious diseases, which are more common in LMIC, can contribute to the occurrence of NCD in endemic countries. Indeed, one of the main drivers of NCD is chronic inflammation. Cardiovascular diseases (CVD), type 2 diabetes, and metabolic syndrome are associated with persistent or chronic inflammation, even low-grade, by inducing or increasing insulin resistance and atherosclerosis [3, 4]. Infectious diseases, either bacterial, viral or parasitic, when repeated or chronic, generally maintain systemic chronic inflammation, which can increase the susceptibility to age–related inflammatory status and chronic immune system activation, both of which are recognized as cardio-metabolic diseases risk factors (CMDRF) [5-8]. Furthermore, alteration of the gut microbiota, which is correlated with cardio-metabolic diseases, can be caused by parasite-induced alteration of the microbial community during chronic infection [9, 10]. Indeed, intestinal protozoa such as *Blastocystis hominis* and *Giardia intestinalis* local inflammatory responses during chronic carriage have shown to be linked to dysbiosis [11, 12].

C-reactive protein (CRP) is a sensitive biomarker of subclinical, low-grade inflammation, it level increases during chronic disorders including metabolic ones in apparently healthy individuals [13]. High-sensitivity CRP (hsCRP) is a measurement of very low levels of plasma CRP, which provides an estimation of body inflammatory status, notably low levels of inflammation. Both biomarkers are associated with an increased risk of atherosclerosis and are recognized as independent risk factors of CVD [14-17]. As an example, high hsCRP concentrations have been shown to be associated with the risk of CVD and to predict the occurrence of myocardial infraction in apparently healthy individuals [15].

In Gabon, a country of sub-Saharan Africa, high prevalences of IPIs are reported with high rates among asymptomatic young people and adults who also have numerous CMDRF [18-20]. Recent surveys highlighted a high frequency of pre-high blood pressure and hyperglycemia among apparently healthy adults from urban and semi-urban settings [20]. Interestingly, although these populations carry the usual CMDRF, they are the most frequent IPI carriers and anemics [21]. Identifying the possible relationship between chronic IPI carriage and systemic inflammation would allow identification of IPIs as possible risk factors for CMD in endemic settings. This would promote an integrated approach of NCD and CD control in co-endemic areas.

The present study assessed the level of CRP and hsCRP as biomarkers of systemic inflammation in uninfected and asymptomatic IPI carriers according to the type of parasitosis and the level of urbanization in Gabon.

## Methods

### Ethical considerations

The study protocol was authorized by the Ministry of Public Health of the Gabonese Republic and approved by the National Ethics Committee (under reference PROT N°002/2020/PR/SG/CNE). This was an observational study. The infected patients received appropriate antihelminthic or antiprotozoal treatment according to the national guidelines. Malaria, HIV and Hepatitis positive individuals were sent to the infectiology unit for appropriate case management and follow-up.

### Study sites and populations

Volunteer participants were enrolled from September 2020 to November 2021 in different settlements of Gabon: Libreville (urban area) and Koula-Moutou and Bitam (both considered to be rural areas). Libreville is the capital of Gabon, with approximately 703,939 inhabitants and comprising almost half of the country’s population (Direction Générale de la Statistique 2015). Koula-Moutou, the main city of the province of Ogooué-lolo (southeastern Gabon), is located 588 km from Libreville, with an estimated population of 25,651 inhabitants. Bitam, the main city of Ntem is located in the north of Gabon, in the province of Woleu-ntem, and comprises approximately 27,923 inhabitants.

The recruitment was performed by the teams of Centre de Recherche en pathogènes Infectieux et Patahologies Associées (CREIPA) in Melen, at the Centre Hospitalier Régional Paul Moukambi of Koula-Moutou (CHRPM) and at the medical center of Bifolossi at Bitam.

Participants were included according to the following criteria: being aged 18 years or older, having resided for at least two years in the study area, absence of any clinical symptoms in the last six months, and providing signed informed consent. No volunteers with a serious health problem or with a history of stroke, cardiovascular diseases (CDV), known chronic infection such as HIV, viral hepatitis infections, tuberculosis, or who had taken an antiparasitic treatment in the last six months were included.

### Procedures

The volunteers underwent malaria, HIV, and hepatitis testing by microscopy-based testing. Those who tested positive were not included in the analysis.

The demographic and socioeconomic data of the participants were recorded on a standardized case report form. Approximately five mL of blood samples were collected by venipuncture for C-reactive protein (CRP), high-sensitivity C-reactive protein (hsCRP) measurements.

For the testing of intestinal parasite infections, sterile clean containers were distributed to each participant with clear instructions for stool collection. Stools may be collected in the morning and immediately returned to the study team. However, some participants who did not provide the samples in the morning were allowed to deliver their stool samples to the study team later in the same day.

### Laboratory analysis

#### Inflammatory biomarkers measurement: C-reactive protein (CRP) and high-sensitivity C-reactive protein (hsCRP)

CRP and hsCRP levels were determined using a GETEIN C1100 point of care test (Getein Biotech, INC., Nanjing, Popular Republic of China), which is based on measurement by indirect immunofluorescence using two antibodies: an anti-human CRP monoclonal antibody coupled to the fluorescence latex located at the sample area and an anti-human CRP monoclonal antibody attached to the test line. The measurements were performed according to the manufacturer’s instructions. The serum CRP concentration was considered to be elevated when the level was higher than or equal to 10 mg/L and for hsCRP for levels higher than 3 mg/L.

#### Diagnosis of intestinal parasitic infections

IPIs were diagnosed by direct microscopic examination and merthiolate-iodine-formaldehyde (MIF) staining which allowed identification of helminth eggs and the cystic and vegetative forms of protozoa as described elsewhere [22]. The Kato-katz technique allowed detection and quantification of STHs and *Schistosoma* spp. on a thick slide equivalent to 41.7 mg of stool (number of eggs/g of feces). Coproculture was used for the identification of *Necator americanus* (hookworm) and *Strongyloides stercoralis* larvae. Samples were considered positive when at least one cyst, one egg, or one larva was detected.

### Statistical analysis

The data were analyzed using SPSS 20.0 software (IBM, New York, USA). The qualitative data are presented as percentages and the quantitative data were checked for the normal distribution before applying the appropriate location indicators: mean ± standard deviation for series that have a normal distribution and median and interquartile range (IQR) otherwise. The corrected Chi-square statistical test and Fischer’s exact test were used for the comparisons between the demographic and socioeconomic characteristics in each area.

Univariate logistic regression was performed to assess the unadjusted association between demographic, socioeconomic characteristics and the presence of IPIs; and between the presence of inflammation and age and the area of residence and the presence and types of IPIs. Multivariate logistic regression was used to determine the independent predictors of IPIs, high CRP and high hsCRP levels; models were adjusted for sociodemographic characteristics, area of residence and IPI for biomarkers levels. The multivariate model was theory-based; thus, all predictor variables were included in the multivariate model irrespective of their statistical significance in the bivariate analysis. Adjusted odds ratio (aOR) and crude odds ratio (cOR) were reported at a 95% confidence interval (CI). All *p* - values are two-tailed, and *p* ≤ 0.05 was considered statistically significant.

### Definitions

- Normal CRP level was below 10 mg/L, for hsCRP it was less than or equal to 3 mg/L.
- Coinfection: presence of at least two different types of parasites (helminth and protozoan).
- Unemployed: participants with no income and no gainful activities.

## Results

### Demographic and socioeconomic characteristics of the study population

A total of 518 participants were included, 64.5% (n = 334) in urban area and 35.5% (n = 184) in rural areas. Women predominated in rural areas (59.8%, n = 110) as well as in the urban area (67.4%, n = 225).

The median age was 35 [27 - 46] years overall, with the inhabitants from rural areas being older than those from the urban area (42 [33 - 50] years vs. 32.0 [26 - 44] years *p* < 0.01). Overall, almost half (48.5%, n = 251) of the participants were aged between 30 and 49 years old; the older ones predominated in rural area population relative to the urban area (29.3%, n = 54 vs 12.3%, n = 41, respectively; *p* < 0.01).

Two-thirds of them (68.0%, n = 352) had attended high school or secondary school, mostly from the urban area, while 19.9% (n = 103) from the rural areas did not have a professional activity. However, those with a primary education level predominated in the rural areas compared to the urban area (50.5%, n = 93 vs. 16.2%, n = 54, respectively), and a high school level predominated in the urban area relative to the rural areas (40.4%, n = 135 vs. 4.3%, n = 8, respectively; *p* < 0.01). Likewise, students predominated in the urban area population compared to the rural areas (36.8%, n = 123 vs. 3.8%, n = 7, respectively; *p* < 0.01), and the unemployment predominated in the rural area population (27.2%, n = 50 vs. 15.9%, n = 53, in the urban areas; *p* < 0.01).

### Prevalence of IPIs and relationship with sociodemographic characteristics

The overall prevalence of asymptomatic IPI was of 29.9% (n = 155). The bivariate analysis showed that IPIs were significantly more frequent in the rural areas, in participants older than 29 years of age, those with primary or secondary school attendances, and the unemployed (Table 1). After, the multivariate analysis, only living in a rural area was found to be a risk factor for IPIs (Table 1). Protozoa (n = 114) were more common than helminths (n = 62) in the participants overall (Fig 1). Helminths–protozoa coinfection was found in 13.5% (n = 21) of the infected participants and was more frequent in rural areas than in the urban area (17.9%, n = 19 vs. 4.1%; n = 2, respectively). Likewise, while the helminth prevalence was high in rural areas, protozoa were more prevalent in the urban site (Fig 1).

**Table 1.** Demographic and socioeconomic characteristics of the population overall and according to the presence of IPIs Table 1 shows the comparison of IPI carrier frequencies by area of residence, gender, age, school attendance and occupation. Comparison was made between IPI carriers for all characteristics to determine the associated risk factors by two- and multi-variable logistic regression tests. All groups with the lowest prevalence of IPIs were selected as a reference. OR: odds ratio.

|  |  | Global | IPIs |  |  |  |  |
| --- | --- | --- | --- | --- | --- | --- | --- |
|  |  | N | N (%) | cOR (95%CI) | p-value | aOR (95%CI) | p-value |
| Area | Urban | 334 | 49 (14.7) | Reference | - | Reference | - |
|  | Rural | 184 | 106 (57.6) | 8.0 (5.2-12.0) | < 0.01 | 6.6 (3.2-13.8) | < 0.01 |
| Gender | Female | 335 | 93 (27.8) | Reference | - | Reference | - |
|  | Male | 183 | 62 (33.9) | 1.3 (0.9-2.0) | 0.15 | 1.3 (0.8-2.0) | 0.32 |
| Age (years) | 18-29 | 172 | 31 (18.0) | Reference | - | Reference | - |
|  | 30-49 | 251 | 86 (34.3) | 2.4 (1.5-3.8) | < 0.01 | 1.7 (0.9-3.4) | 0.10 |
|  | ≥ 50 | 95 | 38 (40.0) | 3.0 (1.7-5.3) | < 0.01 | 1.5 (0.8-3.3) | 0.32 |
| School attendance | High school | 143 | 21 (14.7) | Reference | - | Reference | - |
|  | Secondary | 209 | 62 (29.7) | 2.5 (1.4-4.4) | < 0.01 | 1.2 (0.6-2.3) | 0.60 |
|  | Primary | 147 | 66 (44.9) | 3.7 (1.9-6.9) | < 0.01 | 1.5 (0.7-3.3) | 0.29 |
|  | None | 19 | 6 (31.6) | 2.7 (0.9-7.8) | 0.07 | 0.9 (0.2-3.4) | 0.88 |
| Occupation | Students | 130 | 21 (16.2) | Reference | - | Reference | - |
|  | Workers | 285 | 93 (32.6) | 2.5 (1.5-4.3) | < 0.01 | 0.7 (0.3-1.6) | 0.38 |
|  | Unworkers | 103 | 41 (39.8) | 3.4 (1.9-6.3) | < 0.01 | 0.7 (0.3-1.8) | 0.47 |

**Fig 1.**
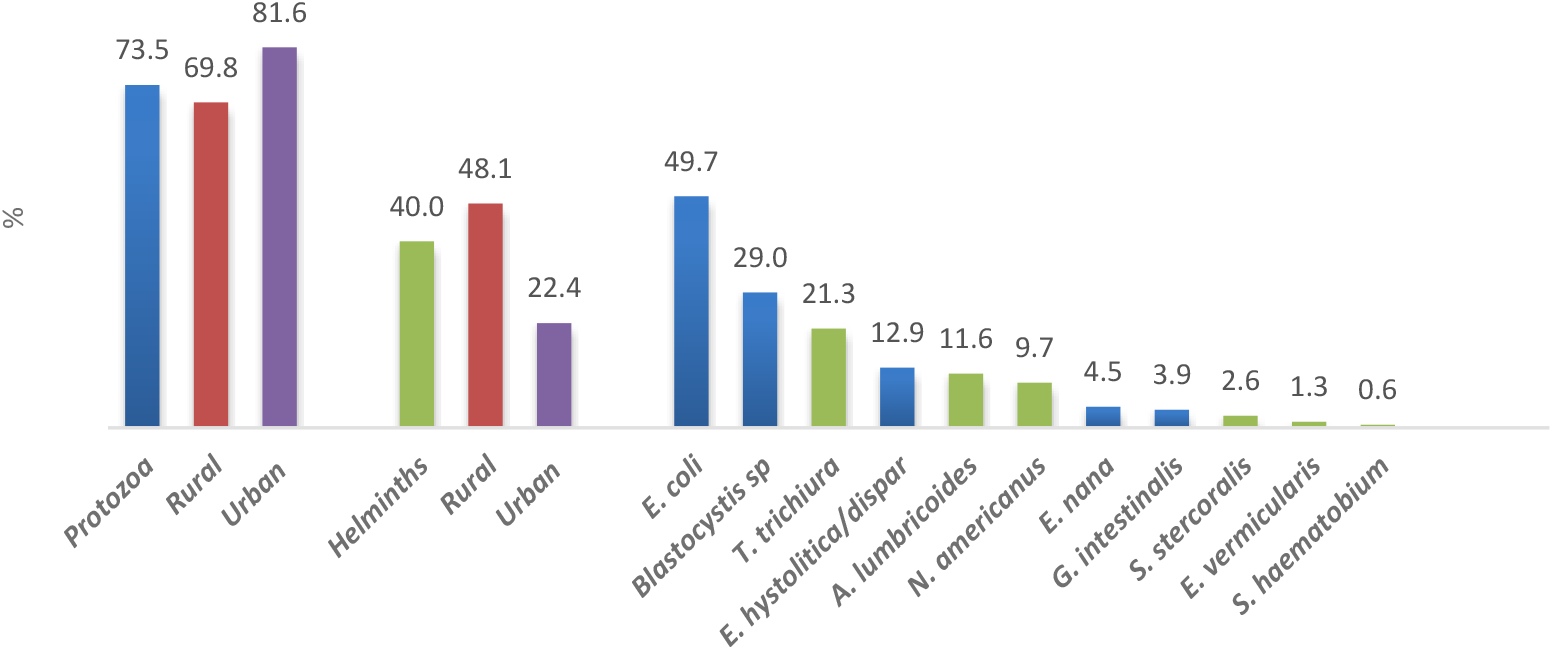
Distribution of intestinal parasite species in the study population. The blue bars of the histogram correspond to the frequency of the protozoa, the green bars to those of the helminths.

Common protozoan parasites were *Entamoeba coli* (n = 77), *Blastocystis* sp (n = 45), and *Entamoeba histolytica/dispar* (n = 20). Common STH included *Trichuris trichiura* (n = 33) and *Ascaris lumbricoides* (n = 18) (Fig 1).

### CRP and hsCRP levels according to the presence or absence of IPI

Overall, the median concentration of hsCRP was of 2.8 [1.4 - 5.2] mg/L and that of CRP was 15.1 [11.5 - 25.0] mg/L. The prevalence of a high hsCRP concentration was 42.5% (n = 220) and that of a high CRP concentration was 15.4% (n = 80). Bivariate analysis showed that a high hsCRP concentration occurred more frequently in inhabitants of rural areas, in older individuals, and in those with IPI. Elevated CRP concentrations predominated in rural inhabitants and in those who were parasitized (Table 2). After adjusting for sociodemographic factors, gender, and sites, only living in a rural area was found to be significantly associated with high hsCRP (aOR 2.0 [CI 1.3-3.1], *p* < 0.01).

**Table 2.** Prevalence of elevated hsCRP and CRP levels according to the sociodemographic characteristics of the population and the results of intestinal parasitosis testing

|  |  | Elevated hsCRP<br>(> 3 mg/L) |  |  | Elevated CRP<br>(≥10 mg/L) |  |  |
| --- | --- | --- | --- | --- | --- | --- | --- |
|  |  | N (%) | cOR 95% CI | p-value | N (%) | cOR 95% CI | p-value |
| Area | Urban | 116 (34.7) | Reference | - | 37 (11.1) | Reference | - |
|  | Rural | 104 (56.5) | 2.4 (1.7-3.5) | < 0.01 | 43 (23.4) | 2.4 (1.8-3.9) | < 0.01 |
| Gender | Female | 135 (40.3) | Reference | - | 49 (14.6) | Reference | - |
|  | Male | 85 (46.4) | 1.3 (0.9-1.9) | 0.18 | 31 (16.9) | 1.2 (0.7-1.9) | 0.49 |
| Age (years) | 18 - 29 | 58 (33.7) | Reference | - | 21 (12.2) | Reference | - |
|  | 30 - 49 | 113 (45.0) | 1.6 (1.1-2.4) | 0.02 | 41 (16.3) | 1.4 (0.8-2.5) | 0.24 |
|  | ≥ 50 | 49 (51.6) | 2.1 (1.3-3.5) | 0.01 | 18 (18.9) | 1.7 (0.8-3.3) | 0.14 |
| IPIs | Absent | 138 (38.0) | Reference | - | 45 (12.4) | Reference | - |
|  | Present | 82 (52.9) | 1.8 (1.3-2.7) | < 0.01 | 35 (22.6) | 2.1 (1.3-3.4) | < 0.01 |
| Type of parasitosis | Protozoa | 44 (47.3) | 1.5 (0.9-2.3) | 0.10 | 17 (18.3) | 1.6 (0.9-2.9) | 0.14 |
|  | STHs | 27 (65.9) | 3.1 (1.6-6.2) | 0.01 | 15 (36.6) | 4.1 (2.0-8.3) | < 0.01 |
|  | **Co-infection** | 11 (52.4) | 1.8 (0.7-4.3) | 0.19 | 3 (14.3) | 1.2 (0.3-4.2) | 0.80 |
| **Number of species** | **1 protozoan** | 31 (45.6) | 13.4 (0.8-2.3) | 0.24 | 14 (20.6) | 1.8 (0.9-3.6) | 0.08 |
|  | **2 protozoa** | 9 (47.4) | 1.5 (0.6-3.7) | 0.42 | 3 (15.8) | 1.3 (0.4-4.7) | 0.67 |
|  | **3 protozoa** | 3 (60.0) | 2.4 (0.4-14.8) | 0.33 | 0 (0.0) | 0.6 (0.03-11.7) | 0.76 |
|  | **4 protozoa** | 1 (100.0) | 4.9 (0.2-120.7) | 0.33 | 0 (0.0) | 2.3 (0.1-58.1) | 0.61 |
|  | **1 STH** | 23 (65.7) | 3.1 (1.5-6.5) | < 0.01 | 14 (40.0) | 4.7 (2.2-9.9) | < 0.01 |
|  | **2 STHs** | 4 (66.7) | 3.3 (0.6-18.0) | 0.18 | 1 (16.7) | 1.4 (0.2-12.4) | 0.76 |
|  | **2 STH/protozoa** | 6 (66.7) | 3.3 (0.8-13.3) | 0.10 | 0 (0.0) | 0.4 (0.02-6.4) | 0.49 |
|  | **3 STH/protozoa** | 2 (22.2) | 0.5 (0.1-2.3) | 0.35 | 1 (11.1) | 0.9 (0.1-7.2) | 0.91 |
| **Protozoan species** | *Blastocystis sp* | 21 (46.7) | 1.4 (0.8-2.7) | 0.26 | 8 (17.8) | 1.5 (0.7-3.5) | 0.31 |
|  | *E. coli* | 34 (43.6) | 1.3 (0.8-2.1) | 0.36 | 11 (14.1) | 1.2 (0.6-2.4) | 0.68 |
|  | *E. histolytica/dispar* | 13 (65.0) | 3.0 (1.2-7.8) | 0.02 | 5 (25.0) | 2.4 (0.8-6.8) | 0.10 |
|  | *E. nana* | 4 (57.1) | 1.2 (0.5-9.9) | 0.31 | 1 (14.3) | 1.2 (0.3-10.0) | 0.88 |
|  | *G. intestinalis* | 6 (100.0) | 21.2 (1.2-378.7) | 0.04 | 1 (16.7) | 1.4 (0.2-12.4) | 0.76 |
| **Helminth species** | *T. trichiura* | 20 (60.6) | 2.5 (1.2-5.2) | 0.01 | 11 (33.3) | 3.5 (1.6-7.8) | < 0.01 |
|  | *A. lumbricoides* | 9 (52.9) | 1.8 (0.7-4.9) | 0.22 | 3 (17.6) | 1.5 (0.4-5.5) | 0.53 |
|  | *E. vermicularis* | 0 (0.0) | 0.3 (0.02-6.8) | 0.47 | 0 (0.0) | 1.4 (0.1-29.6) | 0.83 |
|  | *N. americanus* | 10 (66.7) | 3.3 (1.1-9.7) | 0.03 | 5 (33.3) | 3.5 (1.2-10.8) | 0.03 |
|  | *S. stercoralis* | 4 (100.0) | 14.7 (0.8-274.3) | 0.07 | 1 (25.0) | 2.4 (0.2-23.1) | 0.46 |

This table shows the frequency of elevated CRP and hsCRP concentrations as a function of the areas of residence, the sex, the age, the presence of IPI and according to the different species of parasites. On the other hand, the ORs were calculated by bivariate analysis. STH: soil – transmitted helminth, *E. coli*: *Entamoeba coli*, *E. histolytica/dispar*: *Entamoeba histolytica/dispar*, *E. nana*: *Endolimax nana*, *G. intestinalis*: *Giardia intestinalis*, *T. trichiura*: *Trichuris trichiura*, *A. lumbricoides*: *Ascaris lumbricoides*, *E. vermicularis*: *Enterobius vermicularis*, *N. americanus*: *Necator americanus*, *S. stercoralis*: *Strongyloides stercoralis*

STH carriers were at higher risk of having high hsCRP (aOR 3.1 [CI 1.6 - 6.2], *p* < 0.01) and high CRP (aOR 4.1 [CI 2.0-8.3], *p* < 0.01) levels (Table 2). A trend toward an association between protozoan carriage and high hsCRP (aOR 1.5 [CI 0.3-2.3], *p* = 0.10) and high CRP levels was also observed (Table 2).

All carriers of *Giardia intestinalis* and *Strongyloides stercoralis* had elevated hsCRP levels. However, bivariate analyses indicated that the presence of *Entamoeba histolytica/dispar* and *Giardia intestinalis* were significantly associated with a higher frequency of a high hsCRP concentration compared to the absence of IPI. Participants infected by *Trichuris trichiura* and hookworm had significantly higher hsCRP concentrations (Table 2). Likewise, *Trichuris trichiura* and *Necator americanus* carriages were associated with a high CRP concentrations (Table 2).

### Association between CRP, hsCRP and sociodemographic characteristics in parasitized participants

Male participants tended to be at higher risk of having high hsCRP levels when infected compared to women (Table 3). Living in remote areas was associated with an increased level of CRP. Age did not increase the risk of inflammation in parasitized individuals (Table 3).

**Table 3.** Bivariate and Multivariate analysis of factors associated with high inflammatory biomarkers among the IPI - infected participants

|  |  | Elevated hsCRP |  |  |  |  | Elevated CRP |  |  |  |  |
| --- | --- | --- | --- | --- | --- | --- | --- | --- | --- | --- | --- |
|  |  | N (%) | cOR 95% CI | *p* - value | aOR 95% CI | *p* - value | N (%) | cOR 95% CI | *p* - value | aOR 95% CI | *p* - value |
| **Area** | Urban | 22 (44.9) | Reference | - | Reference | - | 6 (12.2) | Reference | - | Reference | - |
|  | Rural | 60 (56.6) | 1.6 (0.8-3.2) | 0.18 | 0.7 (0.3-1.4) | 0.28 | 29 (27.4) | 2.7 (1.0-7.0) | 0.04 | 2.4 (0.9-6.5) | 0.08 |
| **Gender** | Female | 44 (47.3) | Reference | - | Reference | - | 16 (17.2) | Reference | - | Reference | - |
|  | Male | 38 (61.3) | 1.8 (0.9-3.4) | 0.09 | 1.7 (0.9-3.4) | 0.10 | 19 (30.6) | 2.1 (1.0-4.6) | 1.94 | 2.0 (0.9-4.4) | 0.07 |
| **Age** | 18-29 | 16 (51.6) | Reference | - | Reference | - | 5 (16.1) | Reference | - | Reference | - |
|  | 30-49 | 43 (50.0) | 0.9 (0.4-2.1) | 0.88 | 1.2 (0.5-2.8) | 0.68 | 19 (22.1) | 1.5 (0.5-4.4) | 0.48 | 1.3 (0.4-3.8) | 0.70 |
|  | ≥ 50 | 23 (60.5) | 1.4 (0.5-3.7) | 0.46 | 0.8 (0.3-2.4) | 0.73 | 11 (28.9) | 2.1 (0.6-7.0) | 0.22 | 1.5 (0.4-5.3) | 0.50 |

## Discussion

The present study documented the CRP and hsCRP levels in apparently healthy residents from urban and rural areas of Gabon according to the carriage of intestinal parasites.

The overall prevalence of asymptomatic IPIs found in these populations (29.9%) was lower than in previous reports in other urban and rural areas of Gabon [18, 19]. The higher risk of IPIs, mainly STHs in rural sites, is not surprising. STHs are known to be prevalent in areas where certain existing conditions such as poverty, low level of hygiene and sanitation, and limited access to safe drinking water, favor their spread. The predominance of protozoa compared to STHs in the urban city of Libreville can also be explained by the frequent self-medication with antihelmintics that are freely sold to clients in pharmacies.

Similar to other LMIC, Gabon currently faces the double burden of infectious diseases (ID) and non-communicable diseases (NCD). Indeed, the burden of malaria, HIV, multi-resistant tuberculosis, and other parasite diseases remains high, while increases in NCD morbidity and cardiovascular mortality have also been reported [20, 23]. Compared to high-income countries, high DALYs due to NCD attributed to ID have been reported in LMIC [24]. Thus, while the current westernized lifestyle of inhabitants of SSA urban cities and capitals can partly explain the increase of NCD, this is not the case in rural and remote settlements, where populations live under different conditions with traditional lifestyles.

This study is one of the first to analyze a possible relationship between systemic inflammation, which is a recognized risk factor for CVD, and the presence of IPIs in Central Africa. Our hypothesis that chronic intestinal parasitism can induce a chronic inflammation was tested by measurement of CRP and hsCRP levels, which are biomarkers and indicators of low-grade inflammation in chronic infection [25]. CRP and hsCRP serum levels are independently and significantly associated with the risk of developing ischemic cardiovascular diseases [14]. Moreover, high hsCRP was shown to be associated with the risk of CD and to predict the occurrence of myocardial infarction in healthy individuals [15].

A clear association was found between IPI carriage and biomarkers of systemic inflammation. Consistent with data reported elsewhere, the age of the participants was no found to be a significant risk factor for systemic inflammation [26]. The trends observed in the bivariate analysis is probably due to the predominance of participants who were more than 45 years of age in the rural sites, which are also those in which the highest levels of IPIs were recorded. The significant marked elevation of CRP and hsCRP in these areas, compared to the urban area, can be partly explained by the higher exposure and frequency of STH. These parasites induce mechanical and chemical damage, mucosal inflammation, and host immune responses, which are leading causes of long-term inflammation, thereby representing potential drivers of NCD such as heart, lung, renal, and gastrointestinal diseases as well as cancers [7, 27]. Accordingly, the present results, which highlight an association between chronic pathogenic protozoa and/or STH carriage and increased levels of systemic inflammation biomarkers, suggest that IPIs also contribute to the development of NCD. For instance, some hookworms cause chronic iron-deficiency anemia; and iron deficiency has been reported to be associated with an increase in hsCRP or other inflammatory markers among healthy adults [28, 29]. Through egg migration-induced inflammation, egg embolization, and granuloma development, *Schistosoma* species are causes of cancer, induce renal diseases or pulmonary hypertension which can cause right ventricular failure [30]. Heavy trichuriasis causes chronic infiltration of inflammatory cells in the colon of infected children as well as iron-deficiency anemia [31]. Consistent with this, *Schistosoma* spp., hookworm, and *Trichuris trichiura*, chronic carriage was found associated with increased levels of CRP and hsCRP and thus certainly with low-grade chronic inflammation.

Strongyloidiasis appears to attenuate the systemic inflammation during coinfection or comorbidities [32]. In the present study, asymptomatic *Strongyloides stercoralis* carriers had a high hsCRP levels; further investigations in this regard should be performed.

The absence of an influence of the level of urbanization on the prevalence of participants with high levels of inflammation biomarkers in case of protozoan infection could be explained by the fact that more than 70% of the participants from all the study sites were chronically infected. An association, albeit weak, between the presence of the known pathogens *Entamoeba histolytica* and *Giardia intestinalis* and elevated hsCRP level was found. These pathogenic species, as well *Blastocystis* spp., can cause inflammation [11]. Globally, enteropathogens reduce iron absorption and increase systemic inflammation [8, 33]. The trend toward a higher risk of high biomarker levels in case of helminth-protozoa coinfection confirms the additional impact of protozoa on systemic inflammation.

This study has some limitations. First, although participants were asymptomatic and free of previous antiparasitic treatment, a recent instead of chronic IPI could not be formally excluded. However, the present results already highlight the clear association between the presence of such parasitism and high inflammation biomarker levels. Secondly, the detection of other enteropathogens such as bacteria and fungi, either as invasive pathogens of just colonized ones, was not performed. The analysis of the gut microbiota of the study participants is ongoing. Third, the absence of molecular detection tests for IPIs may have reduced our sample of infected individuals.

Chronic inflammation can be caused by several factors that were not assessed in the present study. However, CRP and hsCRP levels were measured in individuals residing in the same urban or rural city, with comparable living conditions including housing and type of diet, within each site. Thus they should share comparable levels of exposure to some NCD risk factors. Moreover, the analysis performed here are comparable to those in other reports regarding the relationship between infectious diseases or agents and our studied inflammation biomarkers [8, 34]. It is important to note that there is a paucity of previous reports on this topic, especially in adults who carry the double burden of asymptomatic IPI and a higher risk of NCD.

## Conclusion

The present study suggests that STH and pathogenic protozoan intestinal carriage contribute to low-grade inflammation, which is risk factor for NCD, mainly cardiovascular diseases. In rural settlements, permanent exposure to STH may contribute to the increased prevalence of NCD currently observed in Gabon. Although additional information is required to fully confirm our findings, an integrated package of care, including water and sanitation hygiene, as preventive measures against NCD should be emphasized in Gabon.

## Data Availability

All data produced in the study are available upon reasonable request from the authors

## Acknowledgments

The authors would like to thank all the workers of the Centre de Recherche biomédicale En pathogènes Infectieux et Pathologies Associées (CREIPA) and all the technicians of the Unité Mixte de Recherche sur les Agents Infectieux et leur Pathologie (UMRAIP) for their participation to the volunteers recruitment and the sample analysis in Libreville and Bitam. We would like to thank also the technicians of the laboratory of the Centre Hospitalier Régional Paul Moukambi of Koula-Moutou and those of the medical center of Bifolossi at Bitam for their contribution and support in this work, and all the participants to this study.

## Author contributions

**Conceptualization:** Marielle Karine Bouyou Akotet

**Data curation:** Helena Noéline Kono, Mérédith Flore Ada Mengome

**Formal analysis:** Helena Noéline Kono, Cyrille Bisseye

**Funding acquisition:** Marielle Karine Bouyou Akotet

**Investigation:** Helena Noéline Kono, Mérédith Flore Ada Mengome, Bedrich Pongui Ngondza, Roger Hadry Sibi Matotou, Luccheri Ndong Akomezoghe, Bernadette Ekomi, Bridy Chesly Moutombi Ditombi, Jeanne Vanessa Lengongo Koumba, Jacques Mari Ndong Ngomo, Noé Patrick M’Bondoukwé

**Methodology:** Marielle Karine Bouyou Akotet

**Project administration:** Marielle Karine Bouyou Akotet

**Resources:** Marielle Karine Bouyou Akotet

**Supervision:** Denise Patricia Mawili-Mboumba, Marielle Karine Bouyou Akotet

**Validation:** Marielle Karine Bouyou Akotet

**Visualization:** Helena Noéline Kono

**Writing-original draft:** Helena Noéline Kono, Cyrille Bisseye, Denise Patricia Mawili-Mboumba

**Writing-review and editing:** Helena Noéline Kono, Marielle Karine Bouyou Akotet

